## Supplementary Material for "Evaluating the use of social contact data to produce age-specific forecasts of SARS-CoV-2 incidence"

**Supplementary tables**

**Table S1**. Overall scores of all forecasts at all time horizons for both case and infection data.

| **Data type** | **Horizon** | **Model** | **CRPS** | **Bias** | **CRPS relative to “No Interaction” model** |
| --- | --- | --- | --- | --- | --- |
| Cases | 1 Week | CoMix data | 3597.05 | 0.11 | 1.53 |
| Cases | 1 Week | No Contact data | 2404.03 | -0.05 | 1.02 |
| Cases | 1 Week | No Interaction | 2346.04 | -0.16 | 1.00 |
| Cases | 1 Week | Polymod | 8864.86 | 0.16 | 3.78 |
| Cases | 1 Week | Baseline - Fixed value | 4132.47 | 0.00 | 1.76 |
| Cases | 1 Week | Baseline - Exponential | 3156.23 | -0.18 | 1.35 |
| Cases | 2 Weeks | CoMix data | 5888.13 | 0.22 | 1.16 |
| Cases | 2 Weeks | No Contact data | 5017.73 | -0.01 | 0.99 |
| Cases | 2 Weeks | No Interaction | 5088.73 | -0.07 | 1.00 |
| Cases | 2 Weeks | Polymod | 12028.43 | 0.22 | 2.36 |
| Cases | 2 Weeks | Baseline - Fixed value | 6321.75 | -0.09 | 1.24 |
| Cases | 2 Weeks | Baseline - Exponential | 6380.39 | -0.12 | 1.25 |
| Cases | 3 Weeks | CoMix data | 10929.40 | 0.21 | 1.07 |
| Cases | 3 Weeks | No Contact data | 10375.48 | -0.05 | 1.01 |
| Cases | 3 Weeks | No Interaction | 10224.62 | -0.12 | 1.00 |
| Cases | 3 Weeks | Polymod | 17622.45 | 0.17 | 1.72 |
| Cases | 3 Weeks | Baseline - Fixed value | 9542.70 | -0.03 | 0.93 |
| Cases | 3 Weeks | Baseline - Exponential | 11383.50 | -0.12 | 1.11 |
| Cases | 4 Weeks | CoMix data | 16014.57 | 0.22 | 1.01 |
| Cases | 4 Weeks | No Contact data | 16008.11 | 0.00 | 1.01 |
| Cases | 4 Weeks | No Interaction | 15880.62 | -0.10 | 1.00 |
| Cases | 4 Weeks | Polymod | 24365.35 | 0.22 | 1.53 |
| Cases | 4 Weeks | Baseline - Fixed value | 11683.84 | -0.04 | 0.74 |
| Cases | 4 Weeks | Baseline - Exponential | 17269.66 | -0.07 | 1.09 |
| Infections | 1 Week | CoMix data | 10986.46 | -0.02 | 1.05 |
| Infections | 1 Week | No Contact data | 9323.15 | 0.00 | 0.89 |
| Infections | 1 Week | No Interaction | 10470.63 | -0.12 | 1.00 |
| Infections | 1 Week | Polymod | 17315.46 | 0.02 | 1.65 |
| Infections | 1 Week | Baseline - Fixed value | 12645.62 | -0.04 | 1.21 |
| Infections | 1 Week | Baseline - Exponential | 14987.69 | -0.02 | 1.43 |
| Infections | 2 Weeks | CoMix data | 17961.96 | -0.13 | 0.68 |
| Infections | 2 Weeks | No Contact data | 21644.40 | -0.02 | 0.82 |
| Infections | 2 Weeks | No Interaction | 26507.40 | -0.11 | 1.00 |
| Infections | 2 Weeks | Polymod | 26144.07 | -0.03 | 0.99 |
| Infections | 2 Weeks | Baseline - Fixed value | 24051.34 | -0.01 | 0.91 |
| Infections | 2 Weeks | Baseline - Exponential | 37808.06 | 0.00 | 1.43 |
| Infections | 3 Weeks | CoMix data | 27822.94 | -0.16 | 0.64 |
| Infections | 3 Weeks | No Contact data | 37047.29 | -0.04 | 0.85 |
| Infections | 3 Weeks | No Interaction | 43710.42 | -0.11 | 1.00 |
| Infections | 3 Weeks | Polymod | 37724.46 | -0.07 | 0.86 |
| Infections | 3 Weeks | Baseline - Fixed value | 34568.14 | 0.01 | 0.79 |
| Infections | 3 Weeks | Baseline - Exponential | 63242.64 | 0.00 | 1.45 |
| Infections | 4 Weeks | CoMix data | 36953.85 | -0.16 | 0.57 |
| Infections | 4 Weeks | No Contact data | 54826.83 | -0.05 | 0.85 |
| Infections | 4 Weeks | No Interaction | 64820.78 | -0.11 | 1.00 |
| Infections | 4 Weeks | Polymod | 49103.38 | -0.06 | 0.76 |
| Infections | 4 Weeks | Baseline - Fixed value | 44394.54 | -0.06 | 0.68 |
| Infections | 4 Weeks | Baseline - Exponential | 97929.36 | -0.01 | 1.51 |

**Table S2**. Age specific scores of all forecasts at all time horizons for both case and infection data.

| **Data type** | **Horizon** | **Model** | **Age group** | **CRPS relative to “No Interaction” model** | **Bias** |
| --- | --- | --- | --- | --- | --- |
| Infections | 1 Week | CoMix data | 11-15 | 1.37 | -0.15 |
| Infections | 1 Week | No Contact data | 11-15 | 0.89 | -0.13 |
| Infections | 1 Week | No Interaction | 11-15 | 1.00 | -0.11 |
| Infections | 1 Week | Polymod | 11-15 | 1.76 | -0.12 |
| Infections | 1 Week | Baseline - Fixed value | 11-15 | 1.26 | -0.10 |
| Infections | 1 Week | Baseline - Exponential | 11-15 | 1.44 | 0.10 |
| Infections | 2 Weeks | CoMix data | 11-15 | 0.67 | -0.26 |
| Infections | 2 Weeks | No Contact data | 11-15 | 0.75 | 0.00 |
| Infections | 2 Weeks | No Interaction | 11-15 | 1.00 | -0.06 |
| Infections | 2 Weeks | Polymod | 11-15 | 0.95 | -0.14 |
| Infections | 2 Weeks | Baseline - Fixed value | 11-15 | 0.86 | -0.10 |
| Infections | 2 Weeks | Baseline - Exponential | 11-15 | 1.53 | 0.03 |
| Infections | 3 Weeks | CoMix data | 11-15 | 0.53 | -0.19 |
| Infections | 3 Weeks | No Contact data | 11-15 | 0.75 | -0.08 |
| Infections | 3 Weeks | No Interaction | 11-15 | 1.00 | -0.05 |
| Infections | 3 Weeks | Polymod | 11-15 | 0.69 | -0.15 |
| Infections | 3 Weeks | Baseline - Fixed value | 11-15 | 0.66 | -0.10 |
| Infections | 3 Weeks | Baseline - Exponential | 11-15 | 1.67 | 0.10 |
| Infections | 4 Weeks | CoMix data | 11-15 | 0.43 | -0.23 |
| Infections | 4 Weeks | No Contact data | 11-15 | 0.76 | -0.15 |
| Infections | 4 Weeks | No Interaction | 11-15 | 1.00 | -0.09 |
| Infections | 4 Weeks | Polymod | 11-15 | 0.58 | -0.13 |
| Infections | 4 Weeks | Baseline - Fixed value | 11-15 | 0.63 | -0.24 |
| Infections | 4 Weeks | Baseline - Exponential | 11-15 | 1.85 | 0.03 |
| Infections | 1 Week | CoMix data | 16-24 | 1.25 | 0.08 |
| Infections | 1 Week | No Contact data | 16-24 | 1.02 | 0.01 |
| Infections | 1 Week | No Interaction | 16-24 | 1.00 | -0.16 |
| Infections | 1 Week | Polymod | 16-24 | 1.62 | 0.11 |
| Infections | 1 Week | Baseline - Fixed value | 16-24 | 1.31 | 0.03 |
| Infections | 1 Week | Baseline - Exponential | 16-24 | 1.31 | 0.03 |
| Infections | 2 Weeks | CoMix data | 16-24 | 0.90 | -0.04 |
| Infections | 2 Weeks | No Contact data | 16-24 | 1.08 | -0.05 |
| Infections | 2 Weeks | No Interaction | 16-24 | 1.00 | -0.10 |
| Infections | 2 Weeks | Polymod | 16-24 | 1.09 | 0.14 |
| Infections | 2 Weeks | Baseline - Fixed value | 16-24 | 1.11 | 0.03 |
| Infections | 2 Weeks | Baseline - Exponential | 16-24 | 1.31 | 0.03 |
| Infections | 3 Weeks | CoMix data | 16-24 | 0.84 | -0.08 |
| Infections | 3 Weeks | No Contact data | 16-24 | 1.16 | -0.02 |
| Infections | 3 Weeks | No Interaction | 16-24 | 1.00 | -0.08 |
| Infections | 3 Weeks | Polymod | 16-24 | 1.00 | 0.05 |
| Infections | 3 Weeks | Baseline - Fixed value | 16-24 | 0.91 | 0.10 |
| Infections | 3 Weeks | Baseline - Exponential | 16-24 | 1.31 | 0.03 |
| Infections | 4 Weeks | CoMix data | 16-24 | 0.77 | -0.01 |
| Infections | 4 Weeks | No Contact data | 16-24 | 1.17 | 0.02 |
| Infections | 4 Weeks | No Interaction | 16-24 | 1.00 | -0.07 |
| Infections | 4 Weeks | Polymod | 16-24 | 0.90 | 0.07 |
| Infections | 4 Weeks | Baseline - Fixed value | 16-24 | 0.78 | 0.17 |
| Infections | 4 Weeks | Baseline - Exponential | 16-24 | 1.31 | 0.03 |
| Infections | 1 Week | CoMix data | 2-10 | 1.08 | -0.24 |
| Infections | 1 Week | No Contact data | 2-10 | 1.11 | -0.08 |
| Infections | 1 Week | No Interaction | 2-10 | 1.00 | -0.12 |
| Infections | 1 Week | Polymod | 2-10 | 1.52 | -0.14 |
| Infections | 1 Week | Baseline - Fixed value | 2-10 | 1.29 | -0.24 |
| Infections | 1 Week | Baseline - Exponential | 2-10 | 1.48 | -0.03 |
| Infections | 2 Weeks | CoMix data | 2-10 | 0.65 | -0.34 |
| Infections | 2 Weeks | No Contact data | 2-10 | 0.94 | -0.05 |
| Infections | 2 Weeks | No Interaction | 2-10 | 1.00 | -0.12 |
| Infections | 2 Weeks | Polymod | 2-10 | 0.91 | -0.18 |
| Infections | 2 Weeks | Baseline - Fixed value | 2-10 | 0.89 | -0.31 |
| Infections | 2 Weeks | Baseline - Exponential | 2-10 | 1.40 | -0.03 |
| Infections | 3 Weeks | CoMix data | 2-10 | 0.60 | -0.35 |
| Infections | 3 Weeks | No Contact data | 2-10 | 0.91 | -0.09 |
| Infections | 3 Weeks | No Interaction | 2-10 | 1.00 | -0.14 |
| Infections | 3 Weeks | Polymod | 2-10 | 0.86 | -0.19 |
| Infections | 3 Weeks | Baseline - Fixed value | 2-10 | 0.81 | -0.24 |
| Infections | 3 Weeks | Baseline - Exponential | 2-10 | 1.33 | -0.03 |
| Infections | 4 Weeks | CoMix data | 2-10 | 0.46 | -0.40 |
| Infections | 4 Weeks | No Contact data | 2-10 | 0.85 | -0.10 |
| Infections | 4 Weeks | No Interaction | 2-10 | 1.00 | -0.15 |
| Infections | 4 Weeks | Polymod | 2-10 | 0.65 | -0.23 |
| Infections | 4 Weeks | Baseline - Fixed value | 2-10 | 0.62 | -0.17 |
| Infections | 4 Weeks | Baseline - Exponential | 2-10 | 1.38 | -0.10 |
| Infections | 1 Week | CoMix data | 25-34 | 1.42 | 0.07 |
| Infections | 1 Week | No Contact data | 25-34 | 1.00 | 0.02 |
| Infections | 1 Week | No Interaction | 25-34 | 1.00 | -0.18 |
| Infections | 1 Week | Polymod | 25-34 | 2.64 | 0.13 |
| Infections | 1 Week | Baseline - Fixed value | 25-34 | 1.55 | -0.10 |
| Infections | 1 Week | Baseline - Exponential | 25-34 | 1.67 | -0.10 |
| Infections | 2 Weeks | CoMix data | 25-34 | 0.98 | 0.02 |
| Infections | 2 Weeks | No Contact data | 25-34 | 0.99 | 0.01 |
| Infections | 2 Weeks | No Interaction | 25-34 | 1.00 | -0.09 |
| Infections | 2 Weeks | Polymod | 25-34 | 1.48 | 0.10 |
| Infections | 2 Weeks | Baseline - Fixed value | 25-34 | 1.15 | -0.03 |
| Infections | 2 Weeks | Baseline - Exponential | 25-34 | 1.60 | 0.17 |
| Infections | 3 Weeks | CoMix data | 25-34 | 0.94 | 0.01 |
| Infections | 3 Weeks | No Contact data | 25-34 | 1.05 | 0.07 |
| Infections | 3 Weeks | No Interaction | 25-34 | 1.00 | -0.08 |
| Infections | 3 Weeks | Polymod | 25-34 | 1.26 | 0.06 |
| Infections | 3 Weeks | Baseline - Fixed value | 25-34 | 1.02 | 0.17 |
| Infections | 3 Weeks | Baseline - Exponential | 25-34 | 1.62 | 0.10 |
| Infections | 4 Weeks | CoMix data | 25-34 | 0.88 | 0.01 |
| Infections | 4 Weeks | No Contact data | 25-34 | 1.11 | -0.01 |
| Infections | 4 Weeks | No Interaction | 25-34 | 1.00 | -0.14 |
| Infections | 4 Weeks | Polymod | 25-34 | 1.16 | 0.03 |
| Infections | 4 Weeks | Baseline - Fixed value | 25-34 | 0.94 | 0.10 |
| Infections | 4 Weeks | Baseline - Exponential | 25-34 | 1.86 | 0.03 |
| Infections | 1 Week | CoMix data | 35-49 | 0.76 | -0.05 |
| Infections | 1 Week | No Contact data | 35-49 | 0.77 | -0.09 |
| Infections | 1 Week | No Interaction | 35-49 | 1.00 | -0.15 |
| Infections | 1 Week | Polymod | 35-49 | 1.49 | 0.11 |
| Infections | 1 Week | Baseline - Fixed value | 35-49 | 1.03 | -0.03 |
| Infections | 1 Week | Baseline - Exponential | 35-49 | 1.38 | -0.03 |
| Infections | 2 Weeks | CoMix data | 35-49 | 0.58 | -0.17 |
| Infections | 2 Weeks | No Contact data | 35-49 | 0.70 | -0.13 |
| Infections | 2 Weeks | No Interaction | 35-49 | 1.00 | -0.21 |
| Infections | 2 Weeks | Polymod | 35-49 | 0.94 | -0.02 |
| Infections | 2 Weeks | Baseline - Fixed value | 35-49 | 0.83 | 0.10 |
| Infections | 2 Weeks | Baseline - Exponential | 35-49 | 1.39 | -0.10 |
| Infections | 3 Weeks | CoMix data | 35-49 | 0.54 | -0.23 |
| Infections | 3 Weeks | No Contact data | 35-49 | 0.70 | -0.04 |
| Infections | 3 Weeks | No Interaction | 35-49 | 1.00 | -0.14 |
| Infections | 3 Weeks | Polymod | 35-49 | 0.78 | -0.10 |
| Infections | 3 Weeks | Baseline - Fixed value | 35-49 | 0.70 | -0.03 |
| Infections | 3 Weeks | Baseline - Exponential | 35-49 | 1.36 | -0.03 |
| Infections | 4 Weeks | CoMix data | 35-49 | 0.45 | -0.22 |
| Infections | 4 Weeks | No Contact data | 35-49 | 0.64 | -0.07 |
| Infections | 4 Weeks | No Interaction | 35-49 | 1.00 | -0.15 |
| Infections | 4 Weeks | Polymod | 35-49 | 0.65 | -0.09 |
| Infections | 4 Weeks | Baseline - Fixed value | 35-49 | 0.56 | -0.10 |
| Infections | 4 Weeks | Baseline - Exponential | 35-49 | 1.34 | -0.10 |
| Infections | 1 Week | CoMix data | 50-69 | 0.83 | 0.09 |
| Infections | 1 Week | No Contact data | 50-69 | 0.80 | 0.14 |
| Infections | 1 Week | No Interaction | 50-69 | 1.00 | -0.04 |
| Infections | 1 Week | Polymod | 50-69 | 1.74 | 0.08 |
| Infections | 1 Week | Baseline - Fixed value | 50-69 | 1.15 | 0.31 |
| Infections | 1 Week | Baseline - Exponential | 50-69 | 1.38 | 0.03 |
| Infections | 2 Weeks | CoMix data | 50-69 | 0.56 | -0.05 |
| Infections | 2 Weeks | No Contact data | 50-69 | 0.74 | 0.07 |
| Infections | 2 Weeks | No Interaction | 50-69 | 1.00 | -0.03 |
| Infections | 2 Weeks | Polymod | 50-69 | 0.99 | 0.07 |
| Infections | 2 Weeks | Baseline - Fixed value | 50-69 | 0.88 | 0.10 |
| Infections | 2 Weeks | Baseline - Exponential | 50-69 | 1.39 | 0.03 |
| Infections | 3 Weeks | CoMix data | 50-69 | 0.63 | -0.13 |
| Infections | 3 Weeks | No Contact data | 50-69 | 0.84 | -0.03 |
| Infections | 3 Weeks | No Interaction | 50-69 | 1.00 | -0.07 |
| Infections | 3 Weeks | Polymod | 50-69 | 0.95 | 0.00 |
| Infections | 3 Weeks | Baseline - Fixed value | 50-69 | 0.88 | 0.17 |
| Infections | 3 Weeks | Baseline - Exponential | 50-69 | 1.42 | -0.10 |
| Infections | 4 Weeks | CoMix data | 50-69 | 0.66 | -0.14 |
| Infections | 4 Weeks | No Contact data | 50-69 | 0.89 | -0.07 |
| Infections | 4 Weeks | No Interaction | 50-69 | 1.00 | -0.07 |
| Infections | 4 Weeks | Polymod | 50-69 | 0.94 | -0.04 |
| Infections | 4 Weeks | Baseline - Fixed value | 50-69 | 0.84 | -0.17 |
| Infections | 4 Weeks | Baseline - Exponential | 50-69 | 1.42 | 0.03 |
| Infections | 1 Week | CoMix data | 70+ | 0.65 | 0.04 |
| Infections | 1 Week | No Contact data | 70+ | 0.61 | 0.14 |
| Infections | 1 Week | No Interaction | 70+ | 1.00 | -0.05 |
| Infections | 1 Week | Polymod | 70+ | 0.86 | -0.01 |
| Infections | 1 Week | Baseline - Fixed value | 70+ | 0.94 | -0.17 |
| Infections | 1 Week | Baseline - Exponential | 70+ | 1.50 | -0.17 |
| Infections | 2 Weeks | CoMix data | 70+ | 0.41 | -0.09 |
| Infections | 2 Weeks | No Contact data | 70+ | 0.52 | 0.00 |
| Infections | 2 Weeks | No Interaction | 70+ | 1.00 | -0.15 |
| Infections | 2 Weeks | Polymod | 70+ | 0.52 | -0.16 |
| Infections | 2 Weeks | Baseline - Fixed value | 70+ | 0.63 | 0.10 |
| Infections | 2 Weeks | Baseline - Exponential | 70+ | 1.41 | -0.17 |
| Infections | 3 Weeks | CoMix data | 70+ | 0.37 | -0.18 |
| Infections | 3 Weeks | No Contact data | 70+ | 0.51 | -0.08 |
| Infections | 3 Weeks | No Interaction | 70+ | 1.00 | -0.18 |
| Infections | 3 Weeks | Polymod | 70+ | 0.45 | -0.14 |
| Infections | 3 Weeks | Baseline - Fixed value | 70+ | 0.56 | 0.03 |
| Infections | 3 Weeks | Baseline - Exponential | 70+ | 1.51 | -0.10 |
| Infections | 4 Weeks | CoMix data | 70+ | 0.39 | -0.11 |
| Infections | 4 Weeks | No Contact data | 70+ | 0.55 | 0.02 |
| Infections | 4 Weeks | No Interaction | 70+ | 1.00 | -0.11 |
| Infections | 4 Weeks | Polymod | 70+ | 0.45 | -0.02 |
| Infections | 4 Weeks | Baseline - Fixed value | 70+ | 0.51 | -0.03 |
| Infections | 4 Weeks | Baseline - Exponential | 70+ | 1.62 | -0.03 |
| Cases | 1 Week | CoMix data | 0-9 | 1.70 | -0.03 |
| Cases | 1 Week | No Contact data | 0-9 | 1.40 | -0.13 |
| Cases | 1 Week | No Interaction | 0-9 | 1.00 | -0.14 |
| Cases | 1 Week | Polymod | 0-9 | 5.06 | 0.03 |
| Cases | 1 Week | Baseline - Fixed value | 0-9 | 2.09 | -0.31 |
| Cases | 1 Week | Baseline - Exponential | 0-9 | 1.47 | -0.24 |
| Cases | 2 Weeks | CoMix data | 0-9 | 0.91 | 0.08 |
| Cases | 2 Weeks | No Contact data | 0-9 | 1.11 | -0.09 |
| Cases | 2 Weeks | No Interaction | 0-9 | 1.00 | 0.00 |
| Cases | 2 Weeks | Polymod | 0-9 | 2.27 | 0.06 |
| Cases | 2 Weeks | Baseline - Fixed value | 0-9 | 1.21 | -0.38 |
| Cases | 2 Weeks | Baseline - Exponential | 0-9 | 1.33 | -0.03 |
| Cases | 3 Weeks | CoMix data | 0-9 | 0.87 | 0.08 |
| Cases | 3 Weeks | No Contact data | 0-9 | 1.07 | -0.22 |
| Cases | 3 Weeks | No Interaction | 0-9 | 1.00 | -0.11 |
| Cases | 3 Weeks | Polymod | 0-9 | 1.63 | 0.04 |
| Cases | 3 Weeks | Baseline - Fixed value | 0-9 | 0.96 | -0.31 |
| Cases | 3 Weeks | Baseline - Exponential | 0-9 | 1.24 | -0.03 |
| Cases | 4 Weeks | CoMix data | 0-9 | 0.80 | 0.10 |
| Cases | 4 Weeks | No Contact data | 0-9 | 1.00 | -0.11 |
| Cases | 4 Weeks | No Interaction | 0-9 | 1.00 | -0.05 |
| Cases | 4 Weeks | Polymod | 0-9 | 1.42 | 0.12 |
| Cases | 4 Weeks | Baseline - Fixed value | 0-9 | 0.69 | -0.31 |
| Cases | 4 Weeks | Baseline - Exponential | 0-9 | 1.29 | -0.03 |
| Cases | 1 Week | CoMix data | 10-19 | 1.32 | 0.12 |
| Cases | 1 Week | No Contact data | 10-19 | 1.03 | 0.05 |
| Cases | 1 Week | No Interaction | 10-19 | 1.00 | -0.17 |
| Cases | 1 Week | Polymod | 10-19 | 4.46 | 0.12 |
| Cases | 1 Week | Baseline - Fixed value | 10-19 | 1.46 | -0.17 |
| Cases | 1 Week | Baseline - Exponential | 10-19 | 1.36 | -0.24 |
| Cases | 2 Weeks | CoMix data | 10-19 | 0.97 | 0.23 |
| Cases | 2 Weeks | No Contact data | 10-19 | 0.94 | 0.06 |
| Cases | 2 Weeks | No Interaction | 10-19 | 1.00 | -0.05 |
| Cases | 2 Weeks | Polymod | 10-19 | 2.48 | 0.21 |
| Cases | 2 Weeks | Baseline - Fixed value | 10-19 | 0.95 | -0.24 |
| Cases | 2 Weeks | Baseline - Exponential | 10-19 | 1.38 | -0.17 |
| Cases | 3 Weeks | CoMix data | 10-19 | 1.01 | 0.14 |
| Cases | 3 Weeks | No Contact data | 10-19 | 1.01 | -0.13 |
| Cases | 3 Weeks | No Interaction | 10-19 | 1.00 | -0.19 |
| Cases | 3 Weeks | Polymod | 10-19 | 2.02 | 0.13 |
| Cases | 3 Weeks | Baseline - Fixed value | 10-19 | 0.87 | -0.17 |
| Cases | 3 Weeks | Baseline - Exponential | 10-19 | 1.32 | -0.31 |
| Cases | 4 Weeks | CoMix data | 10-19 | 0.86 | 0.15 |
| Cases | 4 Weeks | No Contact data | 10-19 | 0.96 | -0.07 |
| Cases | 4 Weeks | No Interaction | 10-19 | 1.00 | -0.16 |
| Cases | 4 Weeks | Polymod | 10-19 | 1.58 | 0.23 |
| Cases | 4 Weeks | Baseline - Fixed value | 10-19 | 0.60 | -0.17 |
| Cases | 4 Weeks | Baseline - Exponential | 10-19 | 1.25 | -0.17 |
| Cases | 1 Week | CoMix data | 20-29 | 2.46 | 0.15 |
| Cases | 1 Week | No Contact data | 20-29 | 0.94 | -0.02 |
| Cases | 1 Week | No Interaction | 20-29 | 1.00 | -0.14 |
| Cases | 1 Week | Polymod | 20-29 | 5.12 | 0.22 |
| Cases | 1 Week | Baseline - Fixed value | 20-29 | 1.78 | 0.10 |
| Cases | 1 Week | Baseline - Exponential | 20-29 | 1.40 | -0.03 |
| Cases | 2 Weeks | CoMix data | 20-29 | 1.83 | 0.17 |
| Cases | 2 Weeks | No Contact data | 20-29 | 0.98 | -0.03 |
| Cases | 2 Weeks | No Interaction | 20-29 | 1.00 | -0.03 |
| Cases | 2 Weeks | Polymod | 20-29 | 3.05 | 0.23 |
| Cases | 2 Weeks | Baseline - Fixed value | 20-29 | 1.19 | 0.10 |
| Cases | 2 Weeks | Baseline - Exponential | 20-29 | 1.25 | -0.03 |
| Cases | 3 Weeks | CoMix data | 20-29 | 1.49 | 0.18 |
| Cases | 3 Weeks | No Contact data | 20-29 | 0.99 | 0.00 |
| Cases | 3 Weeks | No Interaction | 20-29 | 1.00 | -0.02 |
| Cases | 3 Weeks | Polymod | 20-29 | 2.06 | 0.23 |
| Cases | 3 Weeks | Baseline - Fixed value | 20-29 | 0.96 | 0.10 |
| Cases | 3 Weeks | Baseline - Exponential | 20-29 | 1.02 | 0.10 |
| Cases | 4 Weeks | CoMix data | 20-29 | 1.31 | 0.18 |
| Cases | 4 Weeks | No Contact data | 20-29 | 1.00 | 0.02 |
| Cases | 4 Weeks | No Interaction | 20-29 | 1.00 | -0.05 |
| Cases | 4 Weeks | Polymod | 20-29 | 1.74 | 0.28 |
| Cases | 4 Weeks | Baseline - Fixed value | 20-29 | 0.72 | 0.03 |
| Cases | 4 Weeks | Baseline - Exponential | 20-29 | 1.00 | -0.03 |
| Cases | 1 Week | CoMix data | 30-39 | 1.60 | 0.09 |
| Cases | 1 Week | No Contact data | 30-39 | 1.12 | -0.18 |
| Cases | 1 Week | No Interaction | 30-39 | 1.00 | -0.21 |
| Cases | 1 Week | Polymod | 30-39 | 3.85 | 0.18 |
| Cases | 1 Week | Baseline - Fixed value | 30-39 | 2.02 | 0.03 |
| Cases | 1 Week | Baseline - Exponential | 30-39 | 1.31 | -0.24 |
| Cases | 2 Weeks | CoMix data | 30-39 | 1.22 | 0.19 |
| Cases | 2 Weeks | No Contact data | 30-39 | 0.99 | -0.10 |
| Cases | 2 Weeks | No Interaction | 30-39 | 1.00 | -0.13 |
| Cases | 2 Weeks | Polymod | 30-39 | 2.45 | 0.22 |
| Cases | 2 Weeks | Baseline - Fixed value | 30-39 | 1.35 | 0.03 |
| Cases | 2 Weeks | Baseline - Exponential | 30-39 | 1.16 | -0.24 |
| Cases | 3 Weeks | CoMix data | 30-39 | 1.10 | 0.17 |
| Cases | 3 Weeks | No Contact data | 30-39 | 1.02 | -0.05 |
| Cases | 3 Weeks | No Interaction | 30-39 | 1.00 | -0.16 |
| Cases | 3 Weeks | Polymod | 30-39 | 1.66 | 0.14 |
| Cases | 3 Weeks | Baseline - Fixed value | 30-39 | 0.94 | 0.03 |
| Cases | 3 Weeks | Baseline - Exponential | 30-39 | 1.05 | -0.24 |
| Cases | 4 Weeks | CoMix data | 30-39 | 1.09 | 0.19 |
| Cases | 4 Weeks | No Contact data | 30-39 | 1.02 | 0.01 |
| Cases | 4 Weeks | No Interaction | 30-39 | 1.00 | -0.14 |
| Cases | 4 Weeks | Polymod | 30-39 | 1.59 | 0.18 |
| Cases | 4 Weeks | Baseline - Fixed value | 30-39 | 0.81 | 0.03 |
| Cases | 4 Weeks | Baseline - Exponential | 30-39 | 1.03 | -0.03 |
| Cases | 1 Week | CoMix data | 40-49 | 1.36 | -0.01 |
| Cases | 1 Week | No Contact data | 40-49 | 1.07 | -0.12 |
| Cases | 1 Week | No Interaction | 40-49 | 1.00 | -0.14 |
| Cases | 1 Week | Polymod | 40-49 | 3.31 | 0.08 |
| Cases | 1 Week | Baseline - Fixed value | 40-49 | 2.00 | -0.03 |
| Cases | 1 Week | Baseline - Exponential | 40-49 | 1.33 | -0.17 |
| Cases | 2 Weeks | CoMix data | 40-49 | 1.07 | 0.13 |
| Cases | 2 Weeks | No Contact data | 40-49 | 1.13 | -0.08 |
| Cases | 2 Weeks | No Interaction | 40-49 | 1.00 | -0.07 |
| Cases | 2 Weeks | Polymod | 40-49 | 2.37 | 0.13 |
| Cases | 2 Weeks | Baseline - Fixed value | 40-49 | 1.55 | -0.10 |
| Cases | 2 Weeks | Baseline - Exponential | 40-49 | 1.16 | -0.24 |
| Cases | 3 Weeks | CoMix data | 40-49 | 1.02 | 0.13 |
| Cases | 3 Weeks | No Contact data | 40-49 | 1.13 | -0.10 |
| Cases | 3 Weeks | No Interaction | 40-49 | 1.00 | -0.16 |
| Cases | 3 Weeks | Polymod | 40-49 | 1.75 | 0.08 |
| Cases | 3 Weeks | Baseline - Fixed value | 40-49 | 1.01 | 0.03 |
| Cases | 3 Weeks | Baseline - Exponential | 40-49 | 1.06 | -0.24 |
| Cases | 4 Weeks | CoMix data | 40-49 | 1.03 | 0.14 |
| Cases | 4 Weeks | No Contact data | 40-49 | 1.14 | -0.05 |
| Cases | 4 Weeks | No Interaction | 40-49 | 1.00 | -0.10 |
| Cases | 4 Weeks | Polymod | 40-49 | 1.62 | 0.14 |
| Cases | 4 Weeks | Baseline - Fixed value | 40-49 | 0.83 | 0.03 |
| Cases | 4 Weeks | Baseline - Exponential | 40-49 | 1.04 | -0.10 |
| Cases | 1 Week | CoMix data | 50-59 | 1.10 | 0.12 |
| Cases | 1 Week | No Contact data | 50-59 | 0.98 | -0.04 |
| Cases | 1 Week | No Interaction | 50-59 | 1.00 | -0.14 |
| Cases | 1 Week | Polymod | 50-59 | 2.48 | 0.15 |
| Cases | 1 Week | Baseline - Fixed value | 50-59 | 1.80 | 0.03 |
| Cases | 1 Week | Baseline - Exponential | 50-59 | 1.28 | -0.10 |
| Cases | 2 Weeks | CoMix data | 50-59 | 0.91 | 0.27 |
| Cases | 2 Weeks | No Contact data | 50-59 | 0.99 | -0.03 |
| Cases | 2 Weeks | No Interaction | 50-59 | 1.00 | -0.07 |
| Cases | 2 Weeks | Polymod | 50-59 | 1.96 | 0.23 |
| Cases | 2 Weeks | Baseline - Fixed value | 50-59 | 1.46 | -0.17 |
| Cases | 2 Weeks | Baseline - Exponential | 50-59 | 1.18 | -0.03 |
| Cases | 3 Weeks | CoMix data | 50-59 | 0.87 | 0.21 |
| Cases | 3 Weeks | No Contact data | 50-59 | 1.03 | -0.09 |
| Cases | 3 Weeks | No Interaction | 50-59 | 1.00 | -0.13 |
| Cases | 3 Weeks | Polymod | 50-59 | 1.40 | 0.16 |
| Cases | 3 Weeks | Baseline - Fixed value | 50-59 | 0.93 | 0.10 |
| Cases | 3 Weeks | Baseline - Exponential | 50-59 | 1.02 | -0.03 |
| Cases | 4 Weeks | CoMix data | 50-59 | 0.88 | 0.25 |
| Cases | 4 Weeks | No Contact data | 50-59 | 1.03 | -0.01 |
| Cases | 4 Weeks | No Interaction | 50-59 | 1.00 | -0.07 |
| Cases | 4 Weeks | Polymod | 50-59 | 1.37 | 0.20 |
| Cases | 4 Weeks | Baseline - Fixed value | 50-59 | 0.78 | -0.03 |
| Cases | 4 Weeks | Baseline - Exponential | 50-59 | 1.01 | 0.03 |
| Cases | 1 Week | CoMix data | 60-69 | 0.97 | 0.16 |
| Cases | 1 Week | No Contact data | 60-69 | 1.02 | -0.02 |
| Cases | 1 Week | No Interaction | 60-69 | 1.00 | -0.19 |
| Cases | 1 Week | Polymod | 60-69 | 2.11 | 0.18 |
| Cases | 1 Week | Baseline - Fixed value | 60-69 | 1.72 | 0.10 |
| Cases | 1 Week | Baseline - Exponential | 60-69 | 1.31 | -0.31 |
| Cases | 2 Weeks | CoMix data | 60-69 | 0.83 | 0.29 |
| Cases | 2 Weeks | No Contact data | 60-69 | 0.99 | 0.02 |
| Cases | 2 Weeks | No Interaction | 60-69 | 1.00 | -0.05 |
| Cases | 2 Weeks | Polymod | 60-69 | 1.60 | 0.29 |
| Cases | 2 Weeks | Baseline - Fixed value | 60-69 | 1.44 | 0.03 |
| Cases | 2 Weeks | Baseline - Exponential | 60-69 | 1.22 | -0.10 |
| Cases | 3 Weeks | CoMix data | 60-69 | 0.69 | 0.31 |
| Cases | 3 Weeks | No Contact data | 60-69 | 0.90 | 0.03 |
| Cases | 3 Weeks | No Interaction | 60-69 | 1.00 | -0.10 |
| Cases | 3 Weeks | Polymod | 60-69 | 1.12 | 0.23 |
| Cases | 3 Weeks | Baseline - Fixed value | 60-69 | 0.88 | 0.03 |
| Cases | 3 Weeks | Baseline - Exponential | 60-69 | 1.08 | -0.10 |
| Cases | 4 Weeks | CoMix data | 60-69 | 0.75 | 0.29 |
| Cases | 4 Weeks | No Contact data | 60-69 | 0.92 | 0.07 |
| Cases | 4 Weeks | No Interaction | 60-69 | 1.00 | -0.10 |
| Cases | 4 Weeks | Polymod | 60-69 | 1.13 | 0.27 |
| Cases | 4 Weeks | Baseline - Fixed value | 60-69 | 0.75 | -0.03 |
| Cases | 4 Weeks | Baseline - Exponential | 60-69 | 1.06 | -0.10 |
| Cases | 1 Week | CoMix data | 70+ | 0.90 | 0.26 |
| Cases | 1 Week | No Contact data | 70+ | 0.75 | 0.10 |
| Cases | 1 Week | No Interaction | 70+ | 1.00 | -0.19 |
| Cases | 1 Week | Polymod | 70+ | 1.46 | 0.33 |
| Cases | 1 Week | Baseline - Fixed value | 70+ | 1.35 | 0.24 |
| Cases | 1 Week | Baseline - Exponential | 70+ | 1.28 | -0.10 |
| Cases | 2 Weeks | CoMix data | 70+ | 0.66 | 0.42 |
| Cases | 2 Weeks | No Contact data | 70+ | 0.77 | 0.16 |
| Cases | 2 Weeks | No Interaction | 70+ | 1.00 | -0.12 |
| Cases | 2 Weeks | Polymod | 70+ | 1.15 | 0.39 |
| Cases | 2 Weeks | Baseline - Fixed value | 70+ | 1.14 | -0.03 |
| Cases | 2 Weeks | Baseline - Exponential | 70+ | 1.28 | -0.10 |
| Cases | 3 Weeks | CoMix data | 70+ | 0.66 | 0.45 |
| Cases | 3 Weeks | No Contact data | 70+ | 0.88 | 0.12 |
| Cases | 3 Weeks | No Interaction | 70+ | 1.00 | -0.12 |
| Cases | 3 Weeks | Polymod | 70+ | 1.08 | 0.36 |
| Cases | 3 Weeks | Baseline - Fixed value | 70+ | 0.89 | -0.03 |
| Cases | 3 Weeks | Baseline - Exponential | 70+ | 1.24 | -0.10 |
| Cases | 4 Weeks | CoMix data | 70+ | 0.72 | 0.44 |
| Cases | 4 Weeks | No Contact data | 70+ | 0.93 | 0.14 |
| Cases | 4 Weeks | No Interaction | 70+ | 1.00 | -0.09 |
| Cases | 4 Weeks | Polymod | 70+ | 1.09 | 0.34 |
| Cases | 4 Weeks | Baseline - Fixed value | 70+ | 0.75 | 0.10 |
| Cases | 4 Weeks | Baseline - Exponential | 70+ | 1.17 | -0.10 |

**Table S3**. Period specific scores of all forecasts at all time horizons for both case and infection data.

| **Data type** | **Horizon** | **Model** | **Period** | **CRPS relative to “No Interaction” model** | **Bias** |
| --- | --- | --- | --- | --- | --- |
| Infections | 1 Week | CoMix data | Lockdown 2 | 0.58 | 0.24 |
| Infections | 1 Week | No Contact data | Lockdown 2 | 0.53 | -0.43 |
| Infections | 1 Week | No Interaction | Lockdown 2 | 1.00 | -0.35 |
| Infections | 1 Week | Polymod | Lockdown 2 | 4.49 | 0.92 |
| Infections | 1 Week | Baseline - Fixed value | Lockdown 2 | 0.90 | -0.71 |
| Infections | 1 Week | Baseline - Exponential | Lockdown 2 | 1.30 | -0.14 |
| Infections | 2 Weeks | CoMix data | Lockdown 2 | 0.74 | 0.60 |
| Infections | 2 Weeks | No Contact data | Lockdown 2 | 0.51 | -0.04 |
| Infections | 2 Weeks | No Interaction | Lockdown 2 | 1.00 | -0.04 |
| Infections | 2 Weeks | Polymod | Lockdown 2 | 3.35 | 1.00 |
| Infections | 2 Weeks | Baseline - Fixed value | Lockdown 2 | 0.65 | -0.14 |
| Infections | 2 Weeks | Baseline - Exponential | Lockdown 2 | 1.28 | -0.14 |
| Infections | 3 Weeks | CoMix data | Lockdown 2 | 1.51 | 0.99 |
| Infections | 3 Weeks | No Contact data | Lockdown 2 | 0.63 | 0.77 |
| Infections | 3 Weeks | No Interaction | Lockdown 2 | 1.00 | 0.27 |
| Infections | 3 Weeks | Polymod | Lockdown 2 | 3.90 | 1.00 |
| Infections | 3 Weeks | Baseline - Fixed value | Lockdown 2 | 0.61 | 0.71 |
| Infections | 3 Weeks | Baseline - Exponential | Lockdown 2 | 1.25 | 0.43 |
| Infections | 4 Weeks | CoMix data | Lockdown 2 | 1.88 | 1.00 |
| Infections | 4 Weeks | No Contact data | Lockdown 2 | 0.66 | 0.79 |
| Infections | 4 Weeks | No Interaction | Lockdown 2 | 1.00 | 0.28 |
| Infections | 4 Weeks | Polymod | Lockdown 2 | 4.78 | 1.00 |
| Infections | 4 Weeks | Baseline - Fixed value | Lockdown 2 | 0.58 | 0.71 |
| Infections | 4 Weeks | Baseline - Exponential | Lockdown 2 | 1.29 | 0.43 |
| Infections | 1 Week | CoMix data | Lockdown 2  easing | 1.22 | 0.27 |
| Infections | 1 Week | No Contact data | Lockdown 2  easing | 0.85 | 0.15 |
| Infections | 1 Week | No Interaction | Lockdown 2  easing | 1.00 | 0.05 |
| Infections | 1 Week | Polymod | Lockdown 2  easing | 2.78 | 0.64 |
| Infections | 1 Week | Baseline - Fixed value | Lockdown 2  easing | 1.16 | 0.14 |
| Infections | 1 Week | Baseline - Exponential | Lockdown 2  easing | 1.32 | 0.00 |
| Infections | 2 Weeks | CoMix data | Lockdown 2  easing | 0.90 | 0.17 |
| Infections | 2 Weeks | No Contact data | Lockdown 2  easing | 0.67 | -0.13 |
| Infections | 2 Weeks | No Interaction | Lockdown 2  easing | 1.00 | -0.10 |
| Infections | 2 Weeks | Polymod | Lockdown 2  easing | 1.46 | 0.41 |
| Infections | 2 Weeks | Baseline - Fixed value | Lockdown 2  easing | 0.84 | 0.00 |
| Infections | 2 Weeks | Baseline - Exponential | Lockdown 2  easing | 1.28 | 0.00 |
| Infections | 3 Weeks | CoMix data | Lockdown 2  easing | 1.05 | -0.09 |
| Infections | 3 Weeks | No Contact data | Lockdown 2  easing | 0.89 | -0.61 |
| Infections | 3 Weeks | No Interaction | Lockdown 2  easing | 1.00 | -0.42 |
| Infections | 3 Weeks | Polymod | Lockdown 2  easing | 1.49 | -0.08 |
| Infections | 3 Weeks | Baseline - Fixed value | Lockdown 2  easing | 0.97 | -0.57 |
| Infections | 3 Weeks | Baseline - Exponential | Lockdown 2  easing | 1.20 | -0.43 |
| Infections | 4 Weeks | CoMix data | Lockdown 2  easing | 1.08 | -0.15 |
| Infections | 4 Weeks | No Contact data | Lockdown 2  easing | 1.01 | -0.77 |
| Infections | 4 Weeks | No Interaction | Lockdown 2  easing | 1.00 | -0.54 |
| Infections | 4 Weeks | Polymod | Lockdown 2  easing | 1.33 | -0.15 |
| Infections | 4 Weeks | Baseline - Fixed value | Lockdown 2  easing | 1.06 | -0.71 |
| Infections | 4 Weeks | Baseline - Exponential | Lockdown 2  easing | 1.21 | -0.43 |
| Infections | 1 Week | CoMix data | Christmas + Lockdown 3 | 0.95 | -0.94 |
| Infections | 1 Week | No Contact data | Christmas + Lockdown 3 | 0.72 | -0.75 |
| Infections | 1 Week | No Interaction | Christmas + Lockdown 3 | 1.00 | -0.63 |
| Infections | 1 Week | Polymod | Christmas + Lockdown 3 | 0.96 | -0.95 |
| Infections | 1 Week | Baseline - Fixed value | Christmas + Lockdown 3 | 1.44 | -1.00 |
| Infections | 1 Week | Baseline - Exponential | Christmas + Lockdown 3 | 1.14 | -0.71 |
| Infections | 2 Weeks | CoMix data | Christmas + Lockdown 3 | 0.78 | -0.93 |
| Infections | 2 Weeks | No Contact data | Christmas + Lockdown 3 | 0.69 | -0.44 |
| Infections | 2 Weeks | No Interaction | Christmas + Lockdown 3 | 1.00 | -0.34 |
| Infections | 2 Weeks | Polymod | Christmas + Lockdown 3 | 0.75 | -0.79 |
| Infections | 2 Weeks | Baseline - Fixed value | Christmas + Lockdown 3 | 1.19 | -0.71 |
| Infections | 2 Weeks | Baseline - Exponential | Christmas + Lockdown 3 | 1.19 | -0.14 |
| Infections | 3 Weeks | CoMix data | Christmas + Lockdown 3 | 0.23 | -0.41 |
| Infections | 3 Weeks | No Contact data | Christmas + Lockdown 3 | 0.56 | 0.41 |
| Infections | 3 Weeks | No Interaction | Christmas + Lockdown 3 | 1.00 | 0.13 |
| Infections | 3 Weeks | Polymod | Christmas + Lockdown 3 | 0.25 | -0.30 |
| Infections | 3 Weeks | Baseline - Fixed value | Christmas + Lockdown 3 | 0.76 | -0.43 |
| Infections | 3 Weeks | Baseline - Exponential | Christmas + Lockdown 3 | 1.37 | 0.14 |
| Infections | 4 Weeks | CoMix data | Christmas + Lockdown 3 | 0.13 | -0.17 |
| Infections | 4 Weeks | No Contact data | Christmas + Lockdown 3 | 0.67 | 0.97 |
| Infections | 4 Weeks | No Interaction | Christmas + Lockdown 3 | 1.00 | 0.22 |
| Infections | 4 Weeks | Polymod | Christmas + Lockdown 3 | 0.14 | -0.09 |
| Infections | 4 Weeks | Baseline - Fixed value | Christmas + Lockdown 3 | 0.46 | -0.43 |
| Infections | 4 Weeks | Baseline - Exponential | Christmas + Lockdown 3 | 1.47 | 0.14 |
| Infections | 1 Week | CoMix data | Lockdown 3  easing | 0.68 | -0.05 |
| Infections | 1 Week | No Contact data | Lockdown 3  easing | 0.95 | 0.10 |
| Infections | 1 Week | No Interaction | Lockdown 3  easing | 1.00 | -0.06 |
| Infections | 1 Week | Polymod | Lockdown 3  easing | 0.93 | 0.19 |
| Infections | 1 Week | Baseline - Fixed value | Lockdown 3  easing | 1.27 | 0.52 |
| Infections | 1 Week | Baseline - Exponential | Lockdown 3  easing | 1.61 | -0.05 |
| Infections | 2 Weeks | CoMix data | Lockdown 3  easing | 0.36 | -0.13 |
| Infections | 2 Weeks | No Contact data | Lockdown 3  easing | 0.85 | 0.11 |
| Infections | 2 Weeks | No Interaction | Lockdown 3  easing | 1.00 | -0.09 |
| Infections | 2 Weeks | Polymod | Lockdown 3  easing | 0.64 | 0.25 |
| Infections | 2 Weeks | Baseline - Fixed value | Lockdown 3  easing | 1.10 | 0.57 |
| Infections | 2 Weeks | Baseline - Exponential | Lockdown 3  easing | 1.68 | 0.00 |
| Infections | 3 Weeks | CoMix data | Lockdown 3  easing | 0.28 | -0.20 |
| Infections | 3 Weeks | No Contact data | Lockdown 3  easing | 0.86 | 0.09 |
| Infections | 3 Weeks | No Interaction | Lockdown 3  easing | 1.00 | -0.08 |
| Infections | 3 Weeks | Polymod | Lockdown 3  easing | 0.62 | 0.24 |
| Infections | 3 Weeks | Baseline - Fixed value | Lockdown 3  easing | 1.01 | 0.76 |
| Infections | 3 Weeks | Baseline - Exponential | Lockdown 3  easing | 1.78 | 0.10 |
| Infections | 4 Weeks | CoMix data | Lockdown 3  easing | 0.17 | -0.10 |
| Infections | 4 Weeks | No Contact data | Lockdown 3  easing | 0.92 | 0.18 |
| Infections | 4 Weeks | No Interaction | Lockdown 3  easing | 1.00 | -0.03 |
| Infections | 4 Weeks | Polymod | Lockdown 3  easing | 0.64 | 0.29 |
| Infections | 4 Weeks | Baseline - Fixed value | Lockdown 3  easing | 0.98 | 0.86 |
| Infections | 4 Weeks | Baseline - Exponential | Lockdown 3  easing | 1.88 | 0.10 |
| Infections | 1 Week | CoMix data | Opening Up | 1.15 | -0.01 |
| Infections | 1 Week | No Contact data | Opening Up | 0.91 | 0.04 |
| Infections | 1 Week | No Interaction | Opening Up | 1.00 | -0.10 |
| Infections | 1 Week | Polymod | Opening Up | 1.80 | -0.13 |
| Infections | 1 Week | Baseline - Fixed value | Opening Up | 1.23 | -0.16 |
| Infections | 1 Week | Baseline - Exponential | Opening Up | 1.41 | 0.04 |
| Infections | 2 Weeks | CoMix data | Opening Up | 0.67 | -0.12 |
| Infections | 2 Weeks | No Contact data | Opening Up | 0.77 | 0.05 |
| Infections | 2 Weeks | No Interaction | Opening Up | 1.00 | -0.06 |
| Infections | 2 Weeks | Polymod | Opening Up | 1.00 | -0.17 |
| Infections | 2 Weeks | Baseline - Fixed value | Opening Up | 0.84 | -0.10 |
| Infections | 2 Weeks | Baseline - Exponential | Opening Up | 1.46 | 0.06 |
| Infections | 3 Weeks | CoMix data | Opening Up | 0.53 | -0.14 |
| Infections | 3 Weeks | No Contact data | Opening Up | 0.77 | -0.01 |
| Infections | 3 Weeks | No Interaction | Opening Up | 1.00 | -0.07 |
| Infections | 3 Weeks | Polymod | Opening Up | 0.74 | -0.16 |
| Infections | 3 Weeks | Baseline - Fixed value | Opening Up | 0.63 | -0.14 |
| Infections | 3 Weeks | Baseline - Exponential | Opening Up | 1.53 | 0.04 |
| Infections | 4 Weeks | CoMix data | Opening Up | 0.45 | -0.16 |
| Infections | 4 Weeks | No Contact data | Opening Up | 0.75 | -0.14 |
| Infections | 4 Weeks | No Interaction | Opening Up | 1.00 | -0.12 |
| Infections | 4 Weeks | Polymod | Opening Up | 0.59 | -0.16 |
| Infections | 4 Weeks | Baseline - Fixed value | Opening Up | 0.46 | -0.27 |
| Infections | 4 Weeks | Baseline - Exponential | Opening Up | 1.61 | -0.04 |
| Cases | 1 Week | CoMix data | Lockdown 2 | 1.30 | -0.01 |
| Cases | 1 Week | No Contact data | Lockdown 2 | 0.96 | -0.05 |
| Cases | 1 Week | No Interaction | Lockdown 2 | 1.00 | -0.14 |
| Cases | 1 Week | Polymod | Lockdown 2 | 1.35 | 0.02 |
| Cases | 1 Week | Baseline - Fixed value | Lockdown 2 | 1.08 | -0.14 |
| Cases | 1 Week | Baseline - Exponential | Lockdown 2 | 1.51 | -0.03 |
| Cases | 2 Weeks | CoMix data | Lockdown 2 | 0.78 | -0.27 |
| Cases | 2 Weeks | No Contact data | Lockdown 2 | 0.97 | -0.26 |
| Cases | 2 Weeks | No Interaction | Lockdown 2 | 1.00 | -0.23 |
| Cases | 2 Weeks | Polymod | Lockdown 2 | 0.79 | -0.20 |
| Cases | 2 Weeks | Baseline - Fixed value | Lockdown 2 | 0.88 | -0.31 |
| Cases | 2 Weeks | Baseline - Exponential | Lockdown 2 | 1.35 | -0.14 |
| Cases | 3 Weeks | CoMix data | Lockdown 2 | 0.82 | -0.40 |
| Cases | 3 Weeks | No Contact data | Lockdown 2 | 1.00 | -0.30 |
| Cases | 3 Weeks | No Interaction | Lockdown 2 | 1.00 | -0.24 |
| Cases | 3 Weeks | Polymod | Lockdown 2 | 0.82 | -0.35 |
| Cases | 3 Weeks | Baseline - Fixed value | Lockdown 2 | 0.89 | -0.26 |
| Cases | 3 Weeks | Baseline - Exponential | Lockdown 2 | 1.28 | -0.20 |
| Cases | 4 Weeks | CoMix data | Lockdown 2 | 0.80 | -0.46 |
| Cases | 4 Weeks | No Contact data | Lockdown 2 | 0.97 | -0.17 |
| Cases | 4 Weeks | No Interaction | Lockdown 2 | 1.00 | -0.17 |
| Cases | 4 Weeks | Polymod | Lockdown 2 | 0.80 | -0.38 |
| Cases | 4 Weeks | Baseline - Fixed value | Lockdown 2 | 0.87 | -0.43 |
| Cases | 4 Weeks | Baseline - Exponential | Lockdown 2 | 1.30 | -0.03 |
| Cases | 1 Week | CoMix data | Lockdown 2  easing | 3.88 | 0.47 |
| Cases | 1 Week | No Contact data | Lockdown 2  easing | 0.58 | -0.01 |
| Cases | 1 Week | No Interaction | Lockdown 2  easing | 1.00 | -0.20 |
| Cases | 1 Week | Polymod | Lockdown 2  easing | 24.10 | 0.96 |
| Cases | 1 Week | Baseline - Fixed value | Lockdown 2  easing | 1.75 | -1.00 |
| Cases | 1 Week | Baseline - Exponential | Lockdown 2  easing | 1.17 | -0.75 |
| Cases | 2 Weeks | CoMix data | Lockdown 2  easing | 3.23 | 0.78 |
| Cases | 2 Weeks | No Contact data | Lockdown 2  easing | 0.37 | -0.20 |
| Cases | 2 Weeks | No Interaction | Lockdown 2  easing | 1.00 | -0.24 |
| Cases | 2 Weeks | Polymod | Lockdown 2  easing | 10.65 | 0.96 |
| Cases | 2 Weeks | Baseline - Fixed value | Lockdown 2  easing | 1.72 | -1.00 |
| Cases | 2 Weeks | Baseline - Exponential | Lockdown 2  easing | 1.16 | -0.50 |
| Cases | 3 Weeks | CoMix data | Lockdown 2  easing | 4.03 | 0.99 |
| Cases | 3 Weeks | No Contact data | Lockdown 2  easing | 1.10 | 0.75 |
| Cases | 3 Weeks | No Interaction | Lockdown 2  easing | 1.00 | 0.38 |
| Cases | 3 Weeks | Polymod | Lockdown 2  easing | 8.02 | 1.00 |
| Cases | 3 Weeks | Baseline - Fixed value | Lockdown 2  easing | 0.52 | 0.25 |
| Cases | 3 Weeks | Baseline - Exponential | Lockdown 2  easing | 1.22 | 0.50 |
| Cases | 4 Weeks | CoMix data | Lockdown 2  easing | 3.78 | 1.00 |
| Cases | 4 Weeks | No Contact data | Lockdown 2  easing | 1.31 | 0.99 |
| Cases | 4 Weeks | No Interaction | Lockdown 2  easing | 1.00 | 0.61 |
| Cases | 4 Weeks | Polymod | Lockdown 2  easing | 6.49 | 1.00 |
| Cases | 4 Weeks | Baseline - Fixed value | Lockdown 2  easing | 0.64 | 0.75 |
| Cases | 4 Weeks | Baseline - Exponential | Lockdown 2  easing | 1.16 | 0.75 |
| Cases | 1 Week | CoMix data | Christmas + Lockdown 3 | 0.83 | 0.49 |
| Cases | 1 Week | No Contact data | Christmas + Lockdown 3 | 0.99 | -0.01 |
| Cases | 1 Week | No Interaction | Christmas + Lockdown 3 | 1.00 | 0.00 |
| Cases | 1 Week | Polymod | Christmas + Lockdown 3 | 1.57 | 0.70 |
| Cases | 1 Week | Baseline - Fixed value | Christmas + Lockdown 3 | 0.62 | 0.88 |
| Cases | 1 Week | Baseline - Exponential | Christmas + Lockdown 3 | 1.17 | 0.00 |
| Cases | 2 Weeks | CoMix data | Christmas + Lockdown 3 | 0.74 | 0.22 |
| Cases | 2 Weeks | No Contact data | Christmas + Lockdown 3 | 1.01 | 0.00 |
| Cases | 2 Weeks | No Interaction | Christmas + Lockdown 3 | 1.00 | 0.00 |
| Cases | 2 Weeks | Polymod | Christmas + Lockdown 3 | 1.30 | 0.68 |
| Cases | 2 Weeks | Baseline - Fixed value | Christmas + Lockdown 3 | 0.59 | 0.00 |
| Cases | 2 Weeks | Baseline - Exponential | Christmas + Lockdown 3 | 1.11 | 0.00 |
| Cases | 3 Weeks | CoMix data | Christmas + Lockdown 3 | 0.82 | 0.01 |
| Cases | 3 Weeks | No Contact data | Christmas + Lockdown 3 | 1.02 | 0.00 |
| Cases | 3 Weeks | No Interaction | Christmas + Lockdown 3 | 1.00 | 0.00 |
| Cases | 3 Weeks | Polymod | Christmas + Lockdown 3 | 1.12 | 0.41 |
| Cases | 3 Weeks | Baseline - Fixed value | Christmas + Lockdown 3 | 0.65 | 0.00 |
| Cases | 3 Weeks | Baseline - Exponential | Christmas + Lockdown 3 | 1.09 | 0.00 |
| Cases | 4 Weeks | CoMix data | Christmas + Lockdown 3 | 0.86 | 0.03 |
| Cases | 4 Weeks | No Contact data | Christmas + Lockdown 3 | 1.03 | 0.00 |
| Cases | 4 Weeks | No Interaction | Christmas + Lockdown 3 | 1.00 | 0.00 |
| Cases | 4 Weeks | Polymod | Christmas + Lockdown 3 | 1.41 | 0.47 |
| Cases | 4 Weeks | Baseline - Fixed value | Christmas + Lockdown 3 | 0.60 | -0.13 |
| Cases | 4 Weeks | Baseline - Exponential | Christmas + Lockdown 3 | 1.11 | 0.00 |
| Cases | 1 Week | CoMix data | Lockdown 3  easing | 0.92 | -0.94 |
| Cases | 1 Week | No Contact data | Lockdown 3  easing | 1.03 | -1.00 |
| Cases | 1 Week | No Interaction | Lockdown 3  easing | 1.00 | -1.00 |
| Cases | 1 Week | Polymod | Lockdown 3  easing | 1.15 | -0.94 |
| Cases | 1 Week | Baseline - Fixed value | Lockdown 3  easing | 1.75 | -1.00 |
| Cases | 1 Week | Baseline - Exponential | Lockdown 3  easing | 0.96 | -1.00 |
| Cases | 2 Weeks | CoMix data | Lockdown 3  easing | 0.95 | -0.74 |
| Cases | 2 Weeks | No Contact data | Lockdown 3  easing | 1.10 | -0.56 |
| Cases | 2 Weeks | No Interaction | Lockdown 3  easing | 1.00 | -0.71 |
| Cases | 2 Weeks | Polymod | Lockdown 3  easing | 1.53 | -0.91 |
| Cases | 2 Weeks | Baseline - Fixed value | Lockdown 3  easing | 2.53 | -1.00 |
| Cases | 2 Weeks | Baseline - Exponential | Lockdown 3  easing | 0.94 | -0.75 |
| Cases | 3 Weeks | CoMix data | Lockdown 3  easing | 1.03 | -0.93 |
| Cases | 3 Weeks | No Contact data | Lockdown 3  easing | 1.07 | -0.81 |
| Cases | 3 Weeks | No Interaction | Lockdown 3  easing | 1.00 | -0.79 |
| Cases | 3 Weeks | Polymod | Lockdown 3  easing | 1.51 | -0.99 |
| Cases | 3 Weeks | Baseline - Fixed value | Lockdown 3  easing | 2.07 | -1.00 |
| Cases | 3 Weeks | Baseline - Exponential | Lockdown 3  easing | 0.94 | -0.75 |
| Cases | 4 Weeks | CoMix data | Lockdown 3  easing | 1.08 | -0.82 |
| Cases | 4 Weeks | No Contact data | Lockdown 3  easing | 1.12 | -0.60 |
| Cases | 4 Weeks | No Interaction | Lockdown 3  easing | 1.00 | -0.59 |
| Cases | 4 Weeks | Polymod | Lockdown 3  easing | 1.83 | -0.99 |
| Cases | 4 Weeks | Baseline - Fixed value | Lockdown 3  easing | 2.67 | -1.00 |
| Cases | 4 Weeks | Baseline - Exponential | Lockdown 3  easing | 0.91 | -0.50 |
| Cases | 1 Week | CoMix data | Opening Up | 1.13 | 0.25 |
| Cases | 1 Week | No Contact data | Opening Up | 1.09 | 0.23 |
| Cases | 1 Week | No Interaction | Opening Up | 1.00 | 0.14 |
| Cases | 1 Week | Polymod | Opening Up | 1.85 | 0.41 |
| Cases | 1 Week | Baseline - Fixed value | Opening Up | 1.98 | 0.58 |
| Cases | 1 Week | Baseline - Exponential | Opening Up | 1.57 | 0.29 |
| Cases | 2 Weeks | CoMix data | Opening Up | 0.90 | 0.41 |
| Cases | 2 Weeks | No Contact data | Opening Up | 1.04 | 0.36 |
| Cases | 2 Weeks | No Interaction | Opening Up | 1.00 | 0.23 |
| Cases | 2 Weeks | Polymod | Opening Up | 1.80 | 0.51 |
| Cases | 2 Weeks | Baseline - Fixed value | Opening Up | 1.53 | 0.58 |
| Cases | 2 Weeks | Baseline - Exponential | Opening Up | 1.19 | 0.08 |
| Cases | 3 Weeks | CoMix data | Opening Up | 0.72 | 0.59 |
| Cases | 3 Weeks | No Contact data | Opening Up | 1.06 | 0.45 |
| Cases | 3 Weeks | No Interaction | Opening Up | 1.00 | 0.26 |
| Cases | 3 Weeks | Polymod | Opening Up | 1.42 | 0.67 |
| Cases | 3 Weeks | Baseline - Fixed value | Opening Up | 0.80 | 0.58 |
| Cases | 3 Weeks | Baseline - Exponential | Opening Up | 0.83 | 0.25 |
| Cases | 4 Weeks | CoMix data | Opening Up | 0.68 | 0.60 |
| Cases | 4 Weeks | No Contact data | Opening Up | 1.06 | 0.47 |
| Cases | 4 Weeks | No Interaction | Opening Up | 1.00 | 0.25 |
| Cases | 4 Weeks | Polymod | Opening Up | 1.34 | 0.72 |
| Cases | 4 Weeks | Baseline - Fixed value | Opening Up | 0.53 | 0.71 |
| Cases | 4 Weeks | Baseline - Exponential | Opening Up | 0.74 | 0.38 |

Supplementary Figures

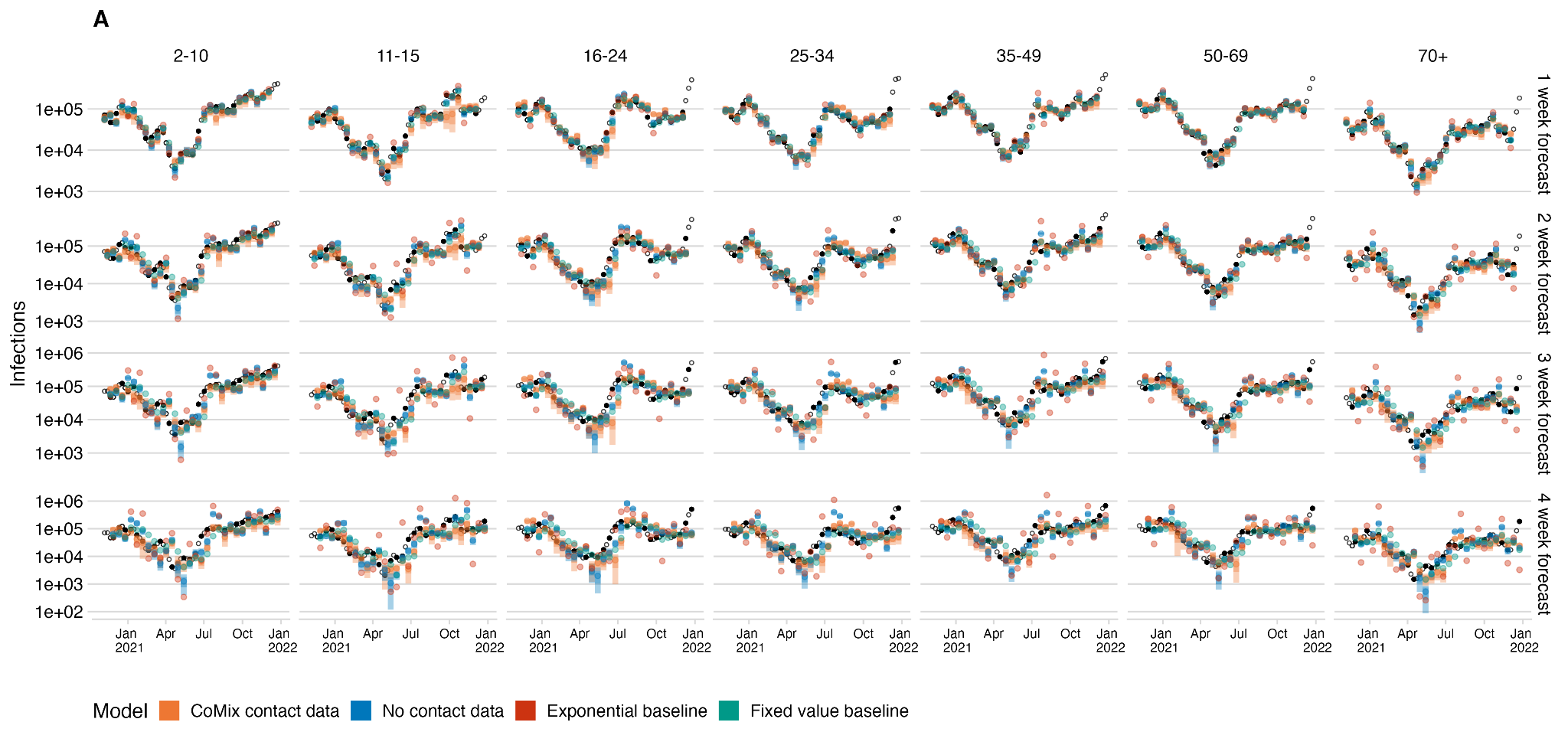

**Figure S1**. Infections forecast using the NGM model with no data alongside the CoMix based next generation model and baseline models for each age group (right to left) and forecast horizon (top to bottom). projected infections from the model (bars) and baselines (coloured points) and black points show infection estimates

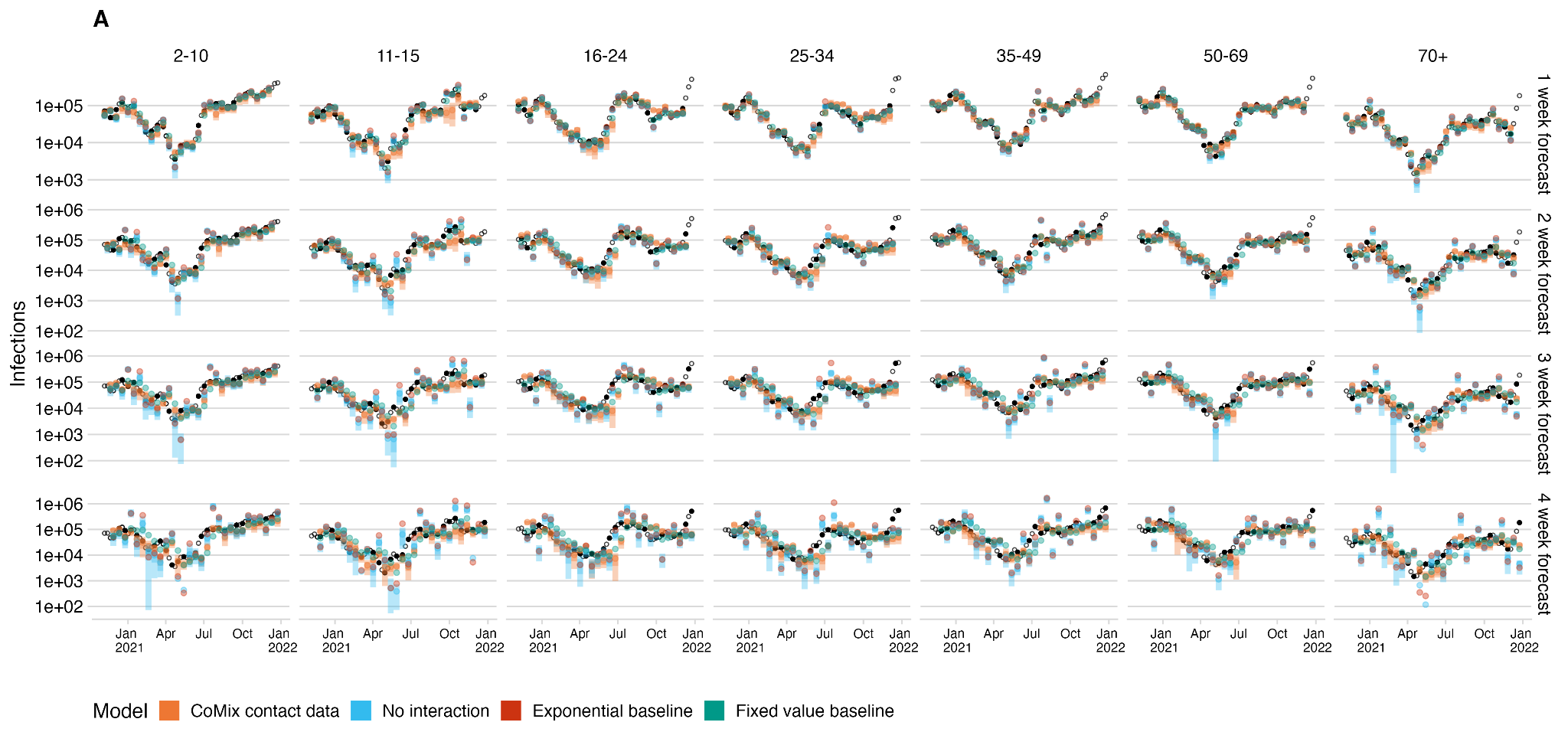

**Figure S2**. Infections forecast using the NGM model with no interaction alongside the CoMix based next generation model and baseline models for each age group (right to left) and forecast horizon (top to bottom). projected infections from the model (bars) and baselines (coloured points) and black points show infection estimates

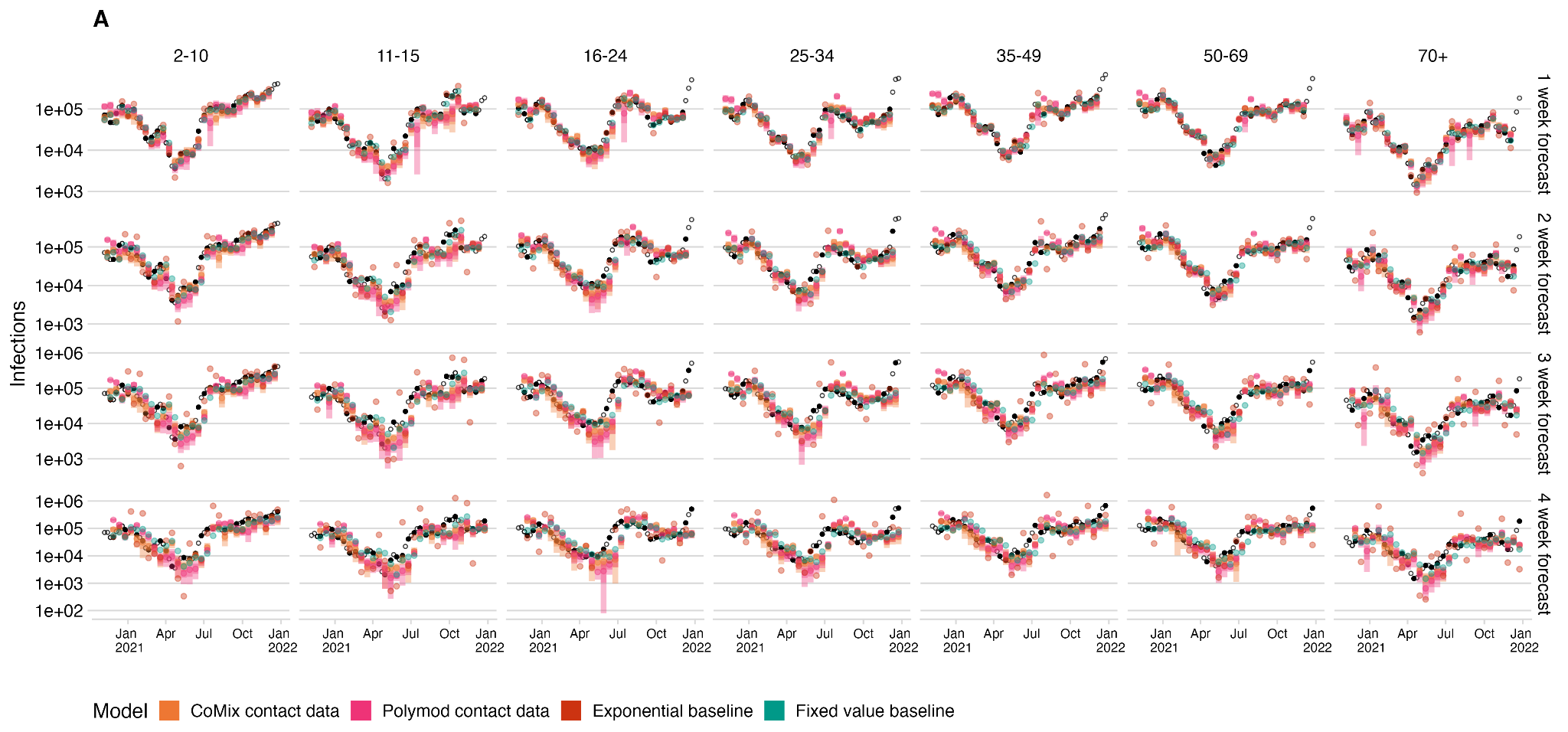

**Figure S3**. Infections forecast using the NGM model using POLYMOD data alongside the CoMix based next generation model and baseline models for each age group (right to left) and forecast horizon (top to bottom). projected infections from the model (bars) and baselines (coloured points) and black points show infection estimates

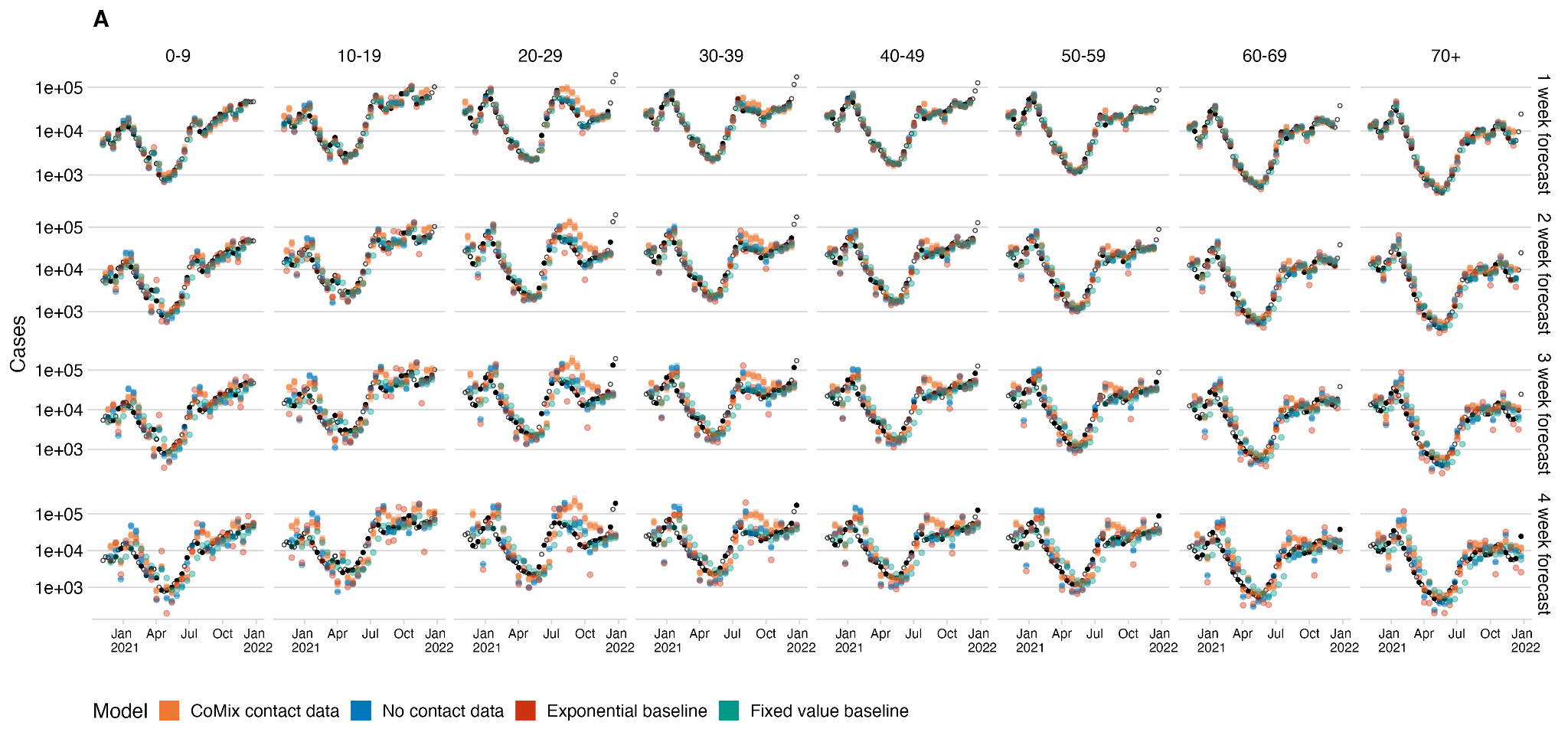

**Figure S4**. Cases forecast using the NGM model with no data alongside the CoMix based next generation model and baseline models for each age group (right to left) and forecast horizon (top to bottom). projected infections from the model (bars) and baselines (coloured points) and black points show cases reported by sequence date

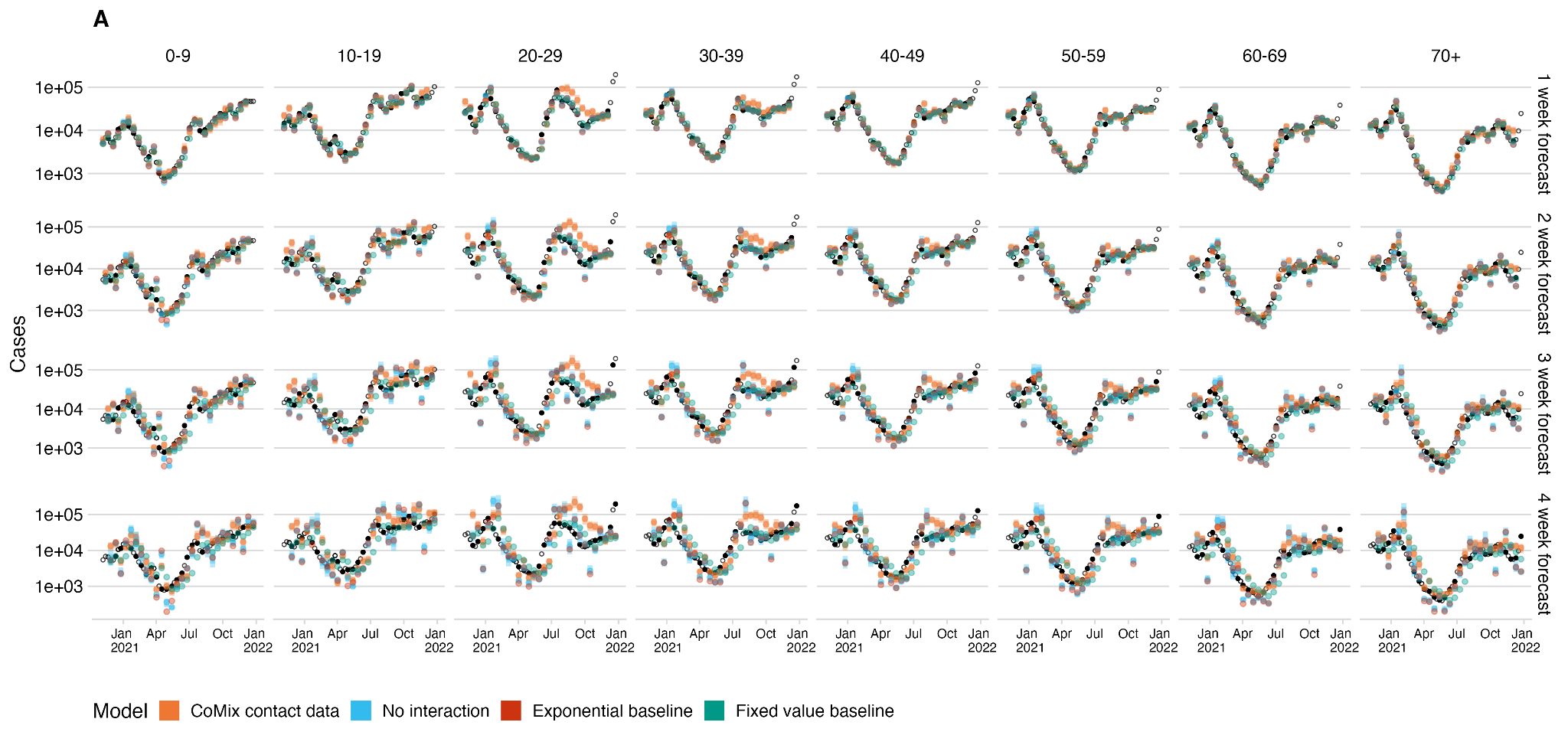

**Figure S5**. Cases forecast using the NGM model with no interaction alongside the CoMix based next generation model and baseline models for each age group (right to left) and forecast horizon (top to bottom). projected infections from the model (bars) and baselines (coloured points) and black points show cases reported by sequence date

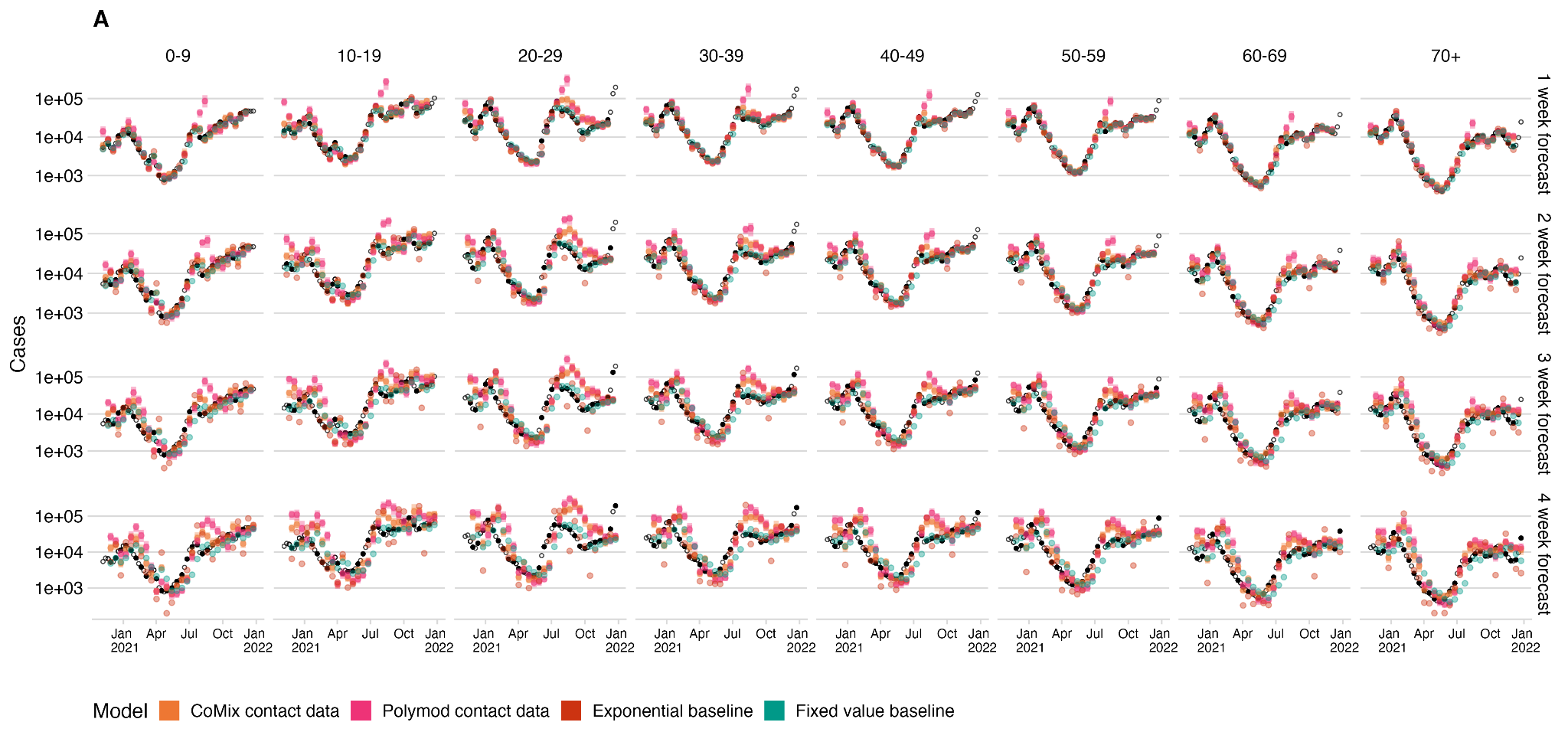

**Figure S6**. Cases forecast using the NGM model using POLYMOD data alongside the CoMix based next generation model and baseline models for each age group (right to left) and forecast horizon (top to bottom). projected infections from the model (bars) and baselines (coloured points) and black points show cases reported by sequence date

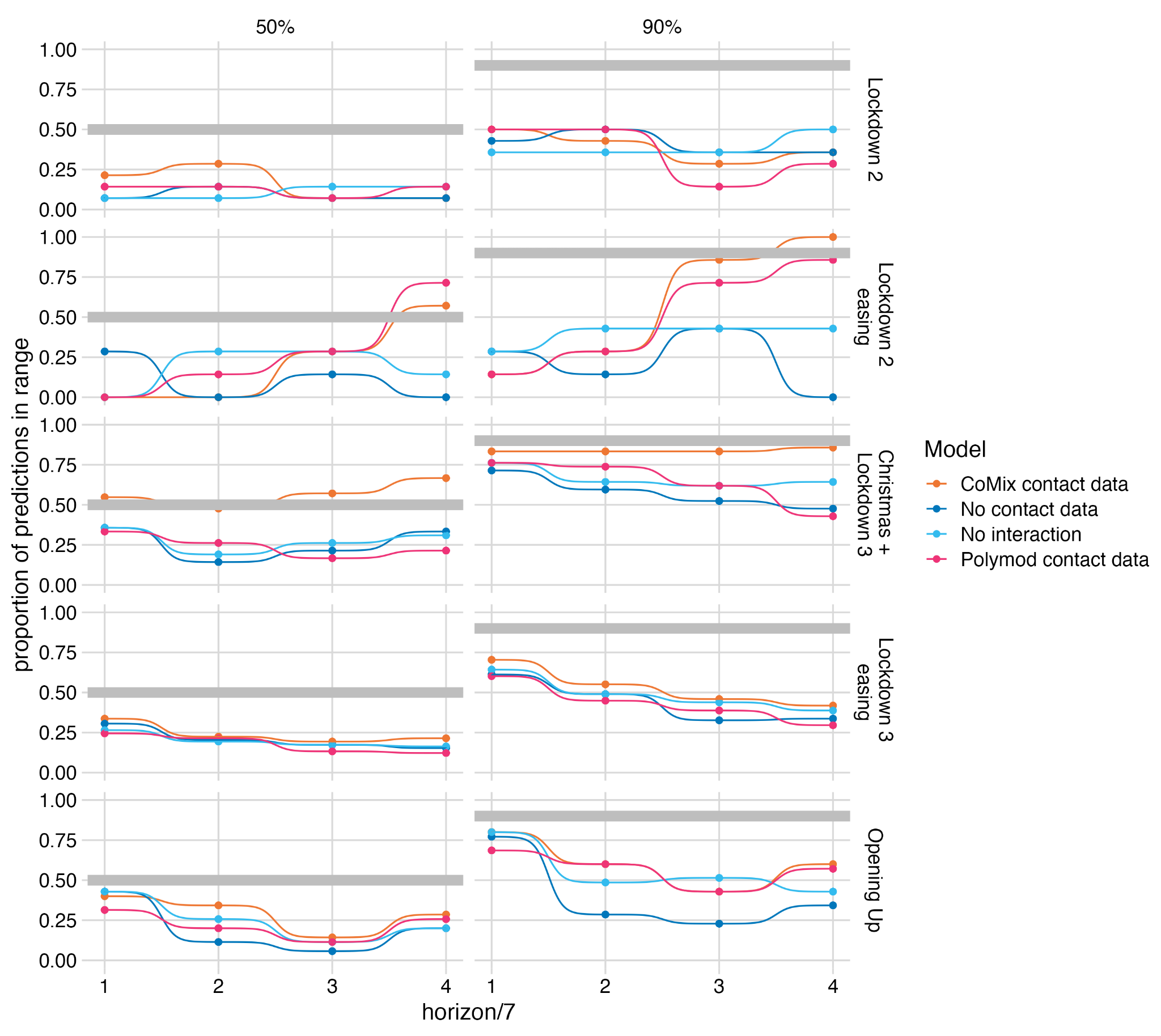

**Figure S7**. The calibration of the forecasts made by each of the next-generation-matrix-based models. The proportion of observed mean incidence of infection estimates (inc2prev) that fall within the 50% and 90% (right and left) central interval of the relevant forecasts of the four models for each pandemic period (top to bottom).

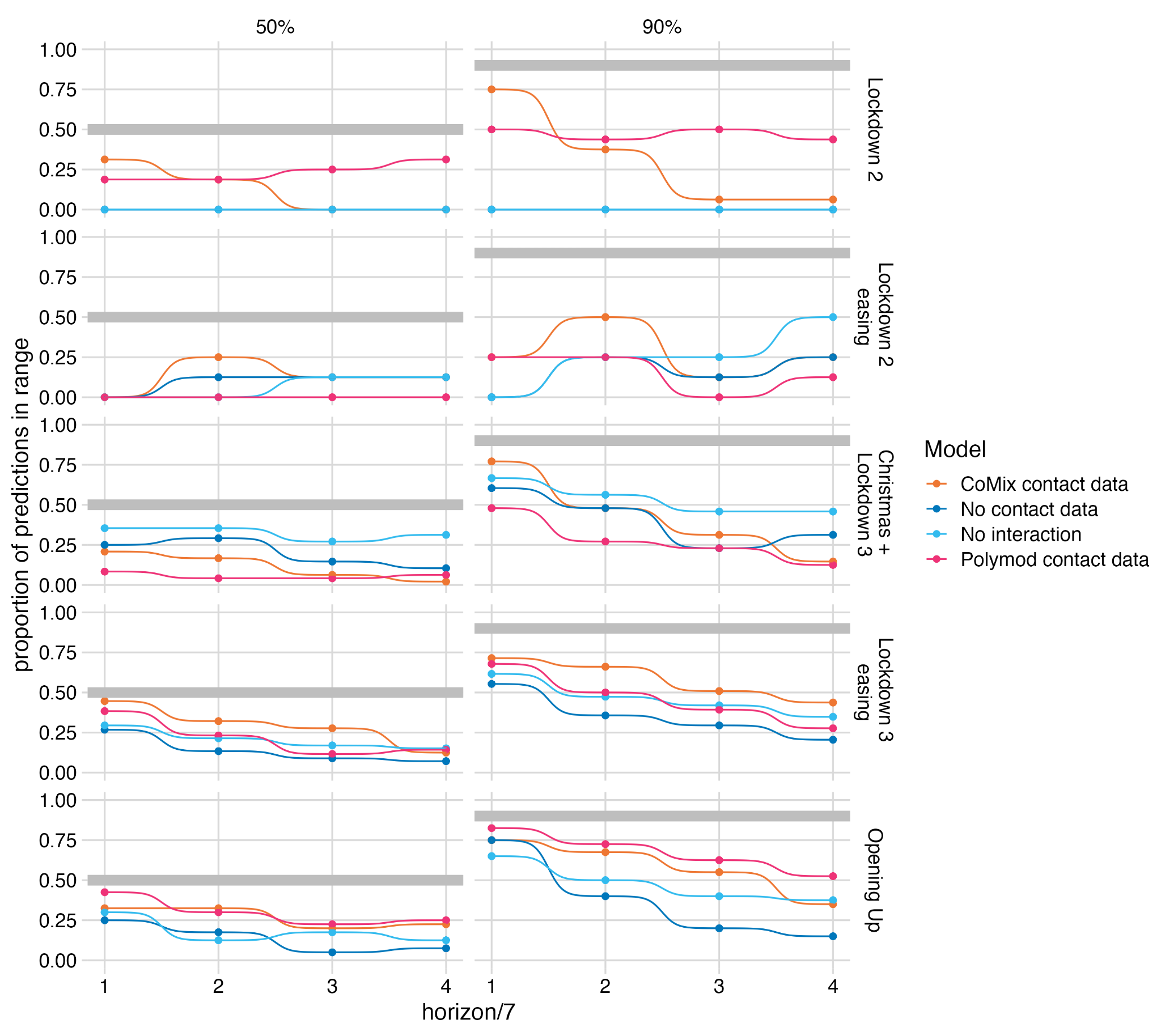

**Figure S8**. The calibration of the forecasts made by each of the next-generation-matrix-based models. The proportion of observed case numbers that fall within the 50% and 90% (right and left) central interval of the relevant forecasts of the four models for each pandemic period (top to bottom).

Supplementary methods

*Model Evaluation*

Scoring rules were implemented using the *scoringutils* package[[1]](https://paperpile.com/c/jFw17n/h4oB). A summary of the methodology, adapted from the *scoringutils* documentation is included below for convenience:

Bias is calculated from predictive Monte-Carlo samples, measured as:

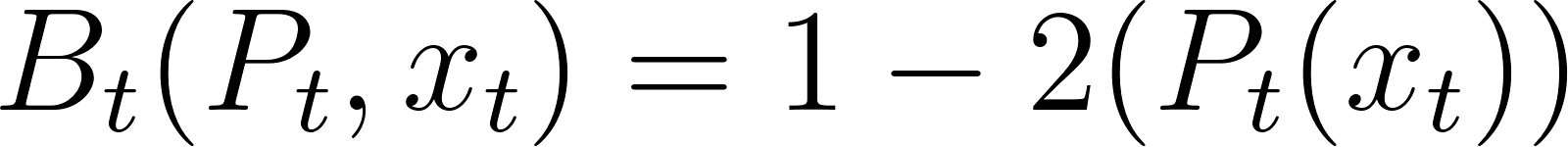
<https://www.codecogs.com/eqnedit.php?latex=B_%7Bt%7D(P_t%2C%20x_t)%20%3D%201-2(P_t(x_t))#0>

where Pt is the empirical cumulative distribution function of the prediction for the true value xt Computationally, Pt(xt) is just calculated as the fraction of predictive samples for xt that are smaller than xt.

The continuous ranked probability score is a proper scoring rule that generalises the absolute error to probabilistic forecasts. It measures the ‘distance’ of the predictive distribution to the observed data-generating distribution. The CRPS is given as

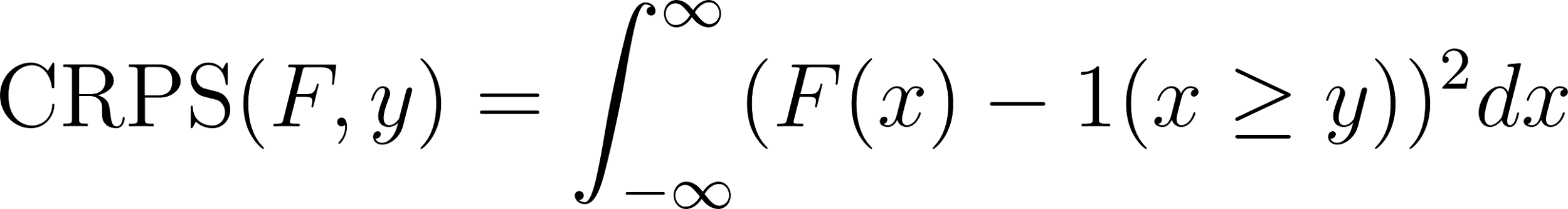
<https://www.codecogs.com/eqnedit.php?latex=%5Ctext%7BCRPS%7D(F%2Cy)%20%3D%20%5Cint_%7B-%5Cinfty%7D%5E%7B%5Cinfty%7D(F(x)%20-%201(x%20%5Cgeq%20y))%5E%7B2%7Ddx#0>

where y is the true observed value and F the CDF of predictive distribution. Often An alternative representation is used:

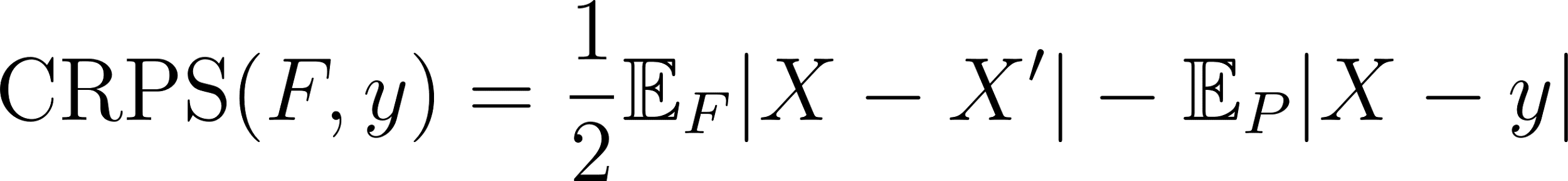
<https://www.codecogs.com/eqnedit.php?latex=%5Ctext%7BCRPS%7D(F%2Cy)%20%3D%20%5Cfrac%7B1%7D%7B2%7D%20%20%5Cmathbb%7BE%7D_%7BF%7D%7CX%20-%20X'%7C%20-%20%5Cmathbb%7BE%7D_%7BP%7D%20%7CX-y%7C#0>

where 𝑋 and 𝑋′ are independent realisations from the predictive distributions 𝐹 with finite first moment and 𝑦 is the true value. In this representation we can simply replace 𝑋 and 𝑋′ by samples sum over all possible combinations to obtain the CRPS.

Smaller values are better. The crps is a good choice for most practical purposes that involve decision making on stationary distributions, as it takes the entire predictive distribution into account. If two forecasters assign the same probability to the true event 𝑦, then the forecaster who assigned high probability to events far away from 𝑦 will still get a worse score. The crps (in contrast to the log score) can at times be quite lenient towards extreme mispredictions. Also, due to it’s similarity to the absolute error, the level of scores depend a lot on the absolute value of what is predicted, which makes it hard to compare scores of forecasts for quantities that are orders of magnitude apart.

1.	Bosse NI, Abbott S, EpiForecasts FS. scoringutils: Utilities for Scoring and Assessing Predictions. 2020.
